# Trends in Repeat Administration of Magnesium for an Exacerbation of Postpartum Preeclampsia: A National Survey

**DOI:** 10.1101/2024.05.21.24307723

**Authors:** Gabriela Dellapiana, Keona Thompson, Kimberly D Gregory

## Abstract

**Background:** Magnesium (Mg) infusion is first-line for seizure prophylaxis in preeclampsia with severe features (PESF). Guidance is lacking regarding utility of an additional course of Mg for disease exacerbation following completion of an initial course. We aimed to determine the national trend among academic institutions for administration of a second course of Mg in patients with an exacerbation of PESF.

**Methods:** A REDCap survey querying response to clinical scenarios related to Mg administration was sent to Maternal-Fetal Medicine program directors (or surrogate if no response) using publicly available contact information. Participants received a $10 incentive.

**Results:** Responses were received from 66 of 95 institutions (69%), representing all 12 ACOG districts. An institutional protocol for postpartum Mg was reported by 70% (N=46) of respondents. Overall, 68% (N=45) would give a second course of Mg for an exacerbation of PESF, with no geographic variation (P=0.969). Most would administer a second course of Mg if the patient had a headache (N=52, 79%) or blood pressure ≥180/120 (N=35, 53%). The decision for a second course of Mg was not influenced by worsening laboratory values, gestational age at delivery, latency from delivery, or maternal demographics.

**Conclusions:** Although there are no standardized guidelines for seizure prophylaxis in postpartum exacerbations of PESF, this national survey suggests that most academic programs favor giving a second course of Mg, especially in patients with headache or severe BP. A randomized trial comparing outcomes in patients who did or did not receive an additional course of Mg is warranted.

## Introduction

Eclampsia is a morbid pregnancy complication characterized by the sudden-onset of a generalized tonic-clonic seizure in a patient with preeclampsia or gestational hypertension. Eclampsia occurs in 2-3% of patients with preeclampsia with severe features (PESF) that goes untreated.^1-3^ Approximately 60% of cases of eclampsia occur antepartum, 20% intrapartum, and 20% postpartum.^4^ Among those cases occurring postpartum, 90% present within the first week of delivery, but eclampsia can manifest up to 6 weeks after birth.^5^

Magnesium (Mg) sulfate infusion is the first-line treatment for the prevention of seizures in patients with PESF, reducing the risk by 58%,^2,6,7^ and the American College of Obstetricians and Gynecologists (ACOG) recommends completing 24 hours of Mg in the immediate postpartum period for most patients.^8^ However, there is no guidance regarding whether or not a patient would benefit from a second course of Mg therapy for seizure prophylaxis in the event of an exacerbation of PESF after completion of the first course.

The purpose of this study is to determine the national trend for the administration or omission of repeat Mg infusion in patients with an exacerbation of PESF by surveying Maternal-Fetal Medicine (MFM) academic providers.

### Study Design

We conducted a voluntary, cross-sectional, web-based, self-administered, nationwide REDCap survey of leaders in perinatology to elicit practice habits regarding repeat administration of Mg. REDCap is a secure web application for managing online surveys. Ethical approval was obtained from the Institutional Review Board. Invited participants included current MFM fellowship directors from all United States-based programs solicited via email using publicly available contact information. If there was no response after 4 mailings, a surrogate program representative was invited using the institutional public website until a response from that institution was received or a maximum of three MFM faculty were approached. If no response was received from a faculty member, a final attempt was made to get a response from an MFM fellow currently at the institution. The fellow was identified from a distribution list of MFM fellows available to one of the investigators. If no response after five individuals had been approached, the program was considered as nonrespondent.

The survey included 16 questions with various clinical scenarios and was designed to be completed in 10 minutes or less. The first page addressed informed consent. Access was limited to those who received a personalized link. Technical functionality of the survey was tested prior to distribution. Respondents were able to review or change answers prior to submission.

Surveys were marked complete if all required questions were answered. Once a completed survey was submitted, edits were no longer available, ensuring a single response per participant. The only demographic data requested was geographic region (as determined by ACOG district; Table 1) and institution (to track response rate). Only completed surveys were included in the analysis. Each survey response was de-identified and assigned a study ID that was exclusively used when sorting and analyzing data. The survey was accessible from March 2022 to June 2022. Participants received a $10 electronic gift card incentive upon completion of the survey.

**Table 1.**
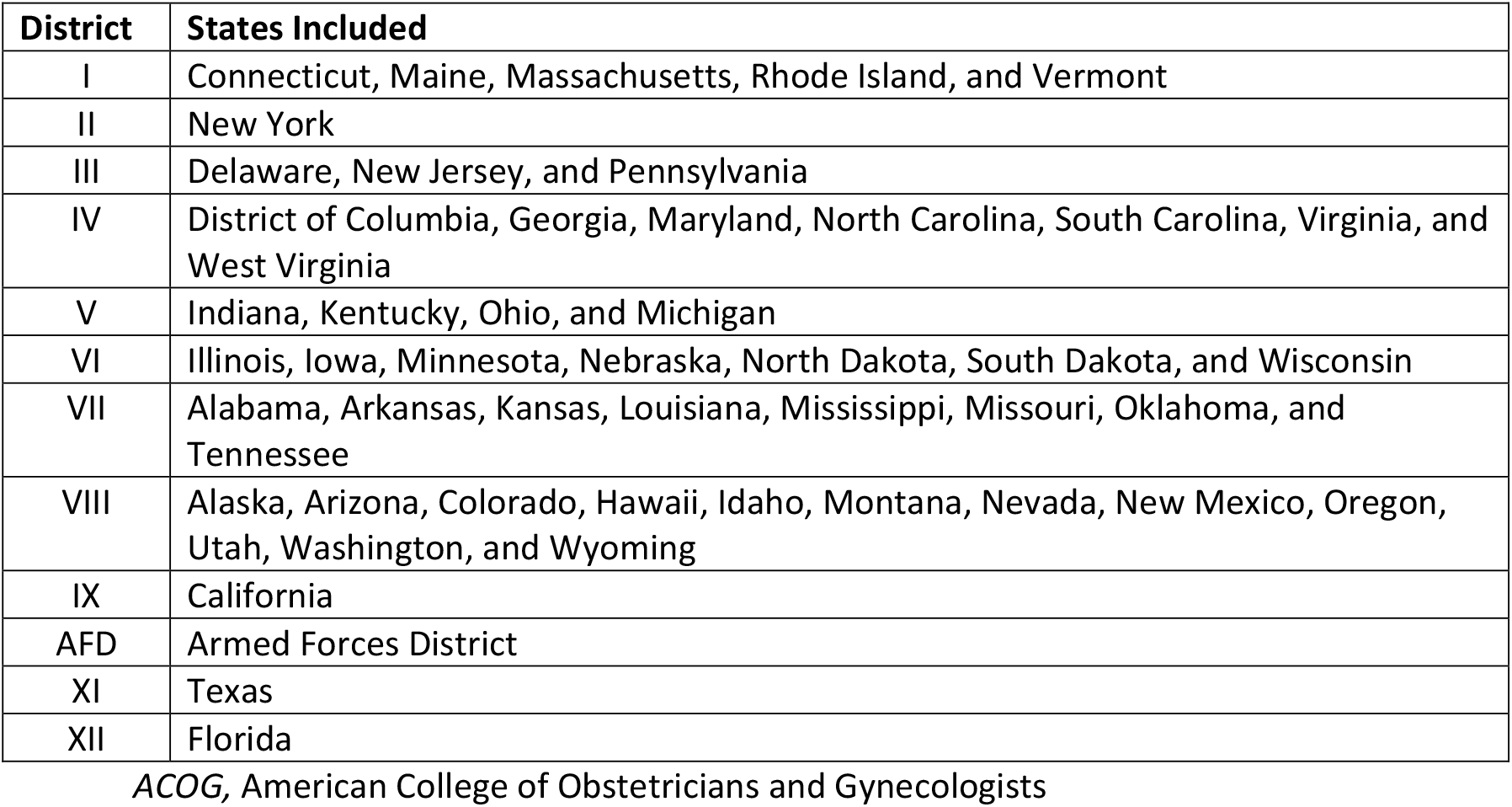
Geographic distribution by ACOG district.

Survey data was analyzed with STATA IC 16.1 (College Station, TX). Data was analyzed using chi-square test and fisher’s exact test as appropriate. Statistical significance was determined by α=0.05.

## Results

Responses were received from 66 of 95 institutions contacted (69% response rate), representing all 12 ACOG districts. MFM program directors comprised 62% of responses (N=41), MFM faculty 24% (N=16), and MFM fellows 14% (N=9). A standard institutional protocol for postpartum Mg was reported by 70% (N=46) of respondents with no difference by geographic region (P=0.666). When asked, “would you give a second course of Mg for seizure prophylaxis for an exacerbation of PESF after completion of the first course,” 68% (N=45) of respondents reported they would (Re-Mg group) and 32% (N=21) reported they would not (No-Mg group). Again, no geographic variation was seen in decision to administer a second course of Mg (P=0.969; Figure 1).

**Figure 1.**
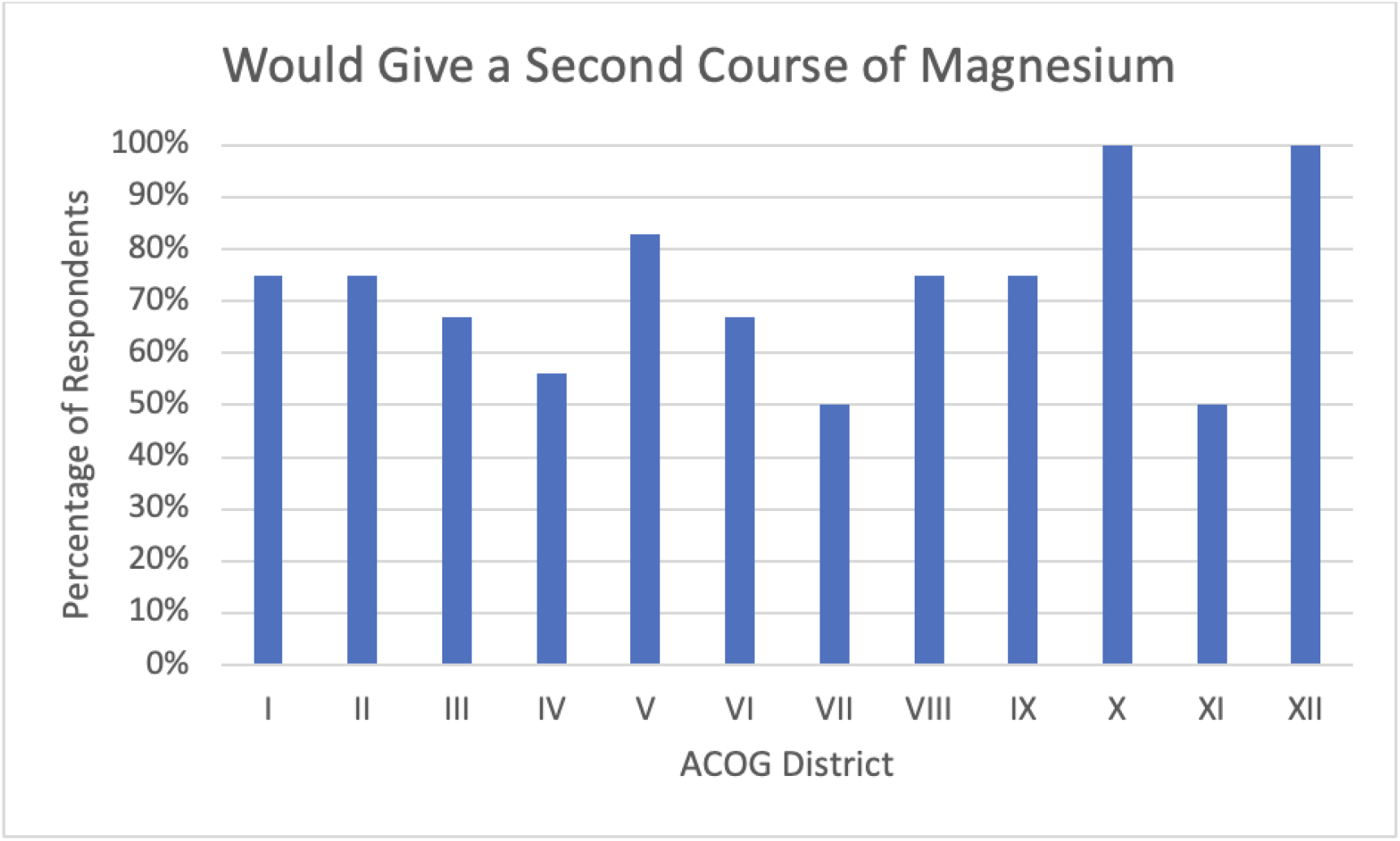
Percent of Maternal-Fetal Medicine physicians who would elect to give a second course of magnesium for an exacerbation of postpartum preeclampsia by ACOG district. *ACOG*, American College of Obstetricians and Gynecologists

The leading clinical scenario in which respondents would choose to administer a second course of Mg was sustained severe hypertension with the coexistence of a headache. For MFMs in the Re-Mg group, presence of a headache was an influencing factor for 98%. Latency from delivery was secondarily influential in that 53% would give a second course of Mg for a headache and hypertension on postpartum day 2, but 96% would on postpartum day 7 and 89% would on postpartum day 14; only 53% would on postpartum week 3-6. Even among MFMs who originally stated they would not give a second course of Mg, the presence of a headache influenced 62% to re-administer Mg with the highest proportion of respondents choosing to do so on postpartum day 2 (38%) or postpartum day 7 (33%).

The next most common reason respondents elected to administer a second course of Mg was based on increasing severity of hypertension (Figure 2). In the Re-Mg group, 47% would give a second course of Mg for systolic blood pressure (SBP) ≥160 and/or diastolic blood pressure (DBP) ≥110 and 62% would for SBP ≥180 and/or DBP ≥120, whereas 36% reported they would give a second course regardless of hypertension severity. Of the respondents who originally stated they would not give a second course, 14% and 33% opted to readminister Mg for SBP≥160 and/or DBP ≥110 and SBP ≥180 and/or DBP ≥110, respectively.

**Figure 2.**
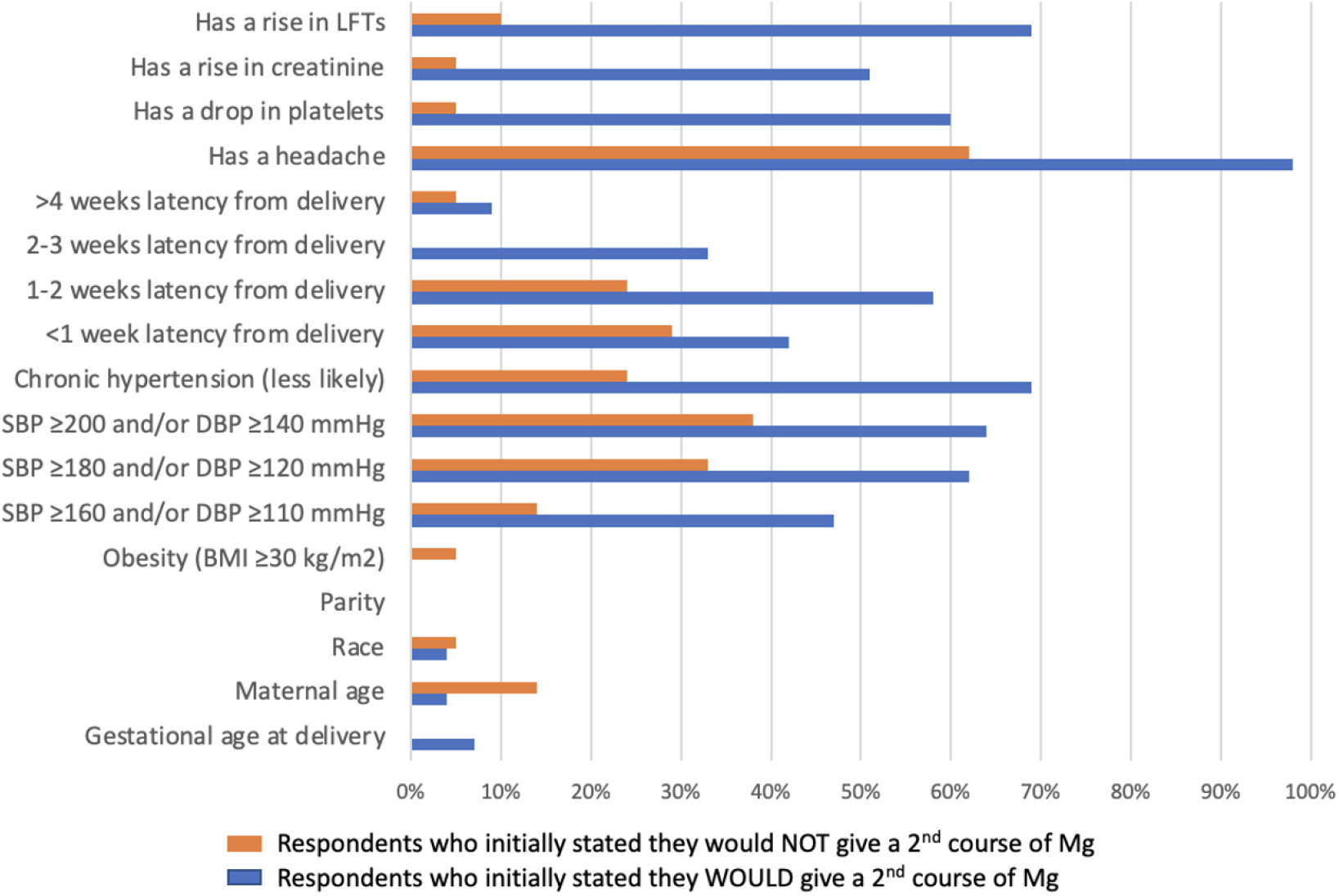
Percentage of Respondents Whose Decision for a 2^nd^ Course of Magnesium for an Exacerbation of PESF would be influenced by: *PESF*, preeclampsia with severe features; *LFTs*, liver function tests; *SBP*, systolic blood pressure; *DBP*, diastolic blood pressure, *BMI*, body mass index

In the Re-Mg group, respondents identified increasing liver function tests (69%), decreasing platelet count (60%), and increasing serum creatinine (51%) as other stimuli for choosing to readminister Mg; changes in laboratory values did not sway the No-Mg group. In both the Re-Mg and No-Mg groups, the decision for a second course of Mg was not influenced by maternal age, race/ethnicity, body mass index, parity, or gestational age at delivery.

## Discussion

Our study explored the practice habits of MFM faculty from academic institutions around the country regarding if and under what circumstances they would administer a second course of Mg for an exacerbation of PESF. Although no standardized guidelines exist, the national trend among academic leaders of obstetric care favors giving a second course of Mg.

Providers queried in our study were most inclined to recommend additional Mg in patients with a headache, corresponding to a previously well-documented risk factor for eclampsia. Systematic reviews have shown that headache precedes seizure activity in up to 80% of patients.^4,9^ Respondents were also more motivated to prescribe a second course of Mg with increasing severity of hypertension. Notably, data has shown that severe hypertension is more strongly correlated with antepartum eclampsia than postpartum eclampsia,^10^ and up to 25% of patients with eclamptic seizure are normotensive immediately preceding convulsions.^11^

We only identified one other study evaluating repeat administration of postpartum Mg. Isler, et al^12^ evaluated 500 patients with preeclampsia and identified 7.6% who received reinstitution of postpartum Mg. In their cohort, patients with chronic hypertension complicated by superimposed preeclampsia were the most likely to receive additional Mg therapy compared to other hypertensive disorders, followed by patients delivered at less than 34 weeks’ gestational age. This is in contrast to our findings; 64% of the Re-Mg group stated that the diagnosis of chronic hypertension would make them *less* likely to readminister Mg, and 34% indicated that chronic hypertension played no role in the decision to repeat prophylaxis. Additionally, 93% of the Re-Mg group and 100% of the No-Mg group indicated that gestational age at delivery had no influence on their choice.

Findings from this study must be evaluated in the context of its limitations. Firstly, we sampled a relatively small sample size compared to the total number of practicing MFM physicians in the country. However, our respondents represent all 12 ACOG districts and are leaders in academic medicine who train obstetric residents and MFM fellows. Studies suggest that providers continue to practice the way they were trained, and that changes in practice can take up to 17 years to become widely implemented despite significant evidence supporting alterations in treatment protocols.^13-14^

Secondly, not all Mg-prescribing obstetrical providers were included in the investigation (e.g., specialist obstetricians, midwives, and family medicine practitioners), but likely they are impacted by interdisciplinary protocols and recommendations from MFM consultants.

### Perspectives

There is a paucity of evidence regarding the need for or efficacy of repeat administration of Mg sulfate for seizure prophylaxis in patients with a postpartum exacerbation of preeclampsia with severe features. That said, a majority of MFM’s recommend a second course of Mg therapy in a patient presenting with headache or progressively severe hypertension. A randomized trial comparing outcomes in patients with an exacerbation of preeclampsia stratified by administration or omission of an additional course of Mg is warranted but unlikely given the large sample size needed to assess for the rare event of eclampsia.

## Data Availability

Data is available from the corresponding author if requested.

## Acknowledgements

Thank you to Samia Saeb, MPH, for assistance with utilizing REDCap.

## Novelty and relevance

1. What is new?
  a. This is the first large-scale survey to our knowledge to ascertain whether academic perinatologists administer or omit an additional magnesium infusion for seizure prophylaxis following completion of the initial course in patients with exacerbation of preeclampsia.
2. What is relevant
  a. Hypertensive disorders of pregnancy are a leading cause of maternal morbidity and mortality
  b. There is currently no guidance for utility or clinical necessity of an additional course of magnesium for seizure prophylaxis
3. Clinical/pathophysiological implications
  a. In the data-free zone that is how to optimally manage an exacerbation of preeclampsia with severe features in the postpartum period, our study provides the national trend among leaders at academic institutions (suggestive of the expert opinion).

